# Knowledge, Attitude and Practice (KAP) study on COVID-19 among the general population of Nepal

**DOI:** 10.1101/2022.09.07.22279527

**Authors:** Salina Shrestha, Rabin Malla, Sadhana Shrestha, Pallavi Singh, Jeevan B. Sherchand

## Abstract

The COVID-19 pandemic has become one of the global health challenges in the current context. In Nepal, the first confirmed case was reported on 23 January 2020, and since then it has resulted in several negative impacts including economic disruption and deterioration of physical and mental health. In such a pandemic, it is indispensable to understand the knowledge and behavioral patterns of the general population regarding COVID-19. Therefore, our study aimed to assess the knowledge, attitude and practice on COVID-19, among the general population and its relationship with socio-demographic conditions.

The questionnaire survey was conducted to collect data from eight districts of Nepal which included Kathmandu, Bhaktapur, Lalitpur, Morang, Sunsari, Rupandehi, Chitwan, and Kaski. Descriptive statistics, parametric and non-parametric statistical tests, and a logistic regression model were used for analysis. The study showed that 93.3% of respondents had knowledge of overall preventive practice whereas only 32% had knowledge of overall symptoms of COVID-19. Regarding attitude, only 14.3% believed that they will get rid of COVID-19 soon. The preventive practice was reduced after lockdown compared to that during lockdown. The respondents with white-collar occupations, high-income, and unmarried were good at KAP. Similarly, highly educated and those residing in urban areas had good knowledge and practice.

The study findings will help in the development of targeted programs to improve the knowledge, attitude, practice of the general population on COVID-19, which is of paramount importance to deal with the existing pandemic and also such possible future waves of the pandemic.

## 1. Introduction

Coronavirus disease 2019 (COVID-19) is an infectious disease first identified in Wuhan, China’s Hubei Province in December 2019 (1). The World Health Organization (WHO) declared this disease a Public Health Emergency of International concern on January 30, 2020, and later declared it a pandemic on March 11, 2020 (2). Now, it has been reported in almost all the countries worldwide. Nepal first confirmed the case of COVID-19 on 23 January 2020 (3).

Prevention and control of the spread of COVID-19 is still a challenge in the current context of emerging new variants of severe acute respiratory syndrome coronavirus −2 (SARS-CoV-2). Though a vaccine has been developed which can reduce the transmission and risk of severity of the disease but cannot stop the transmission. Moreover, the spread of disease is highly dependent on the habit of practicing preventive measures such as personal hygiene, social distancing, use of masks, etc. Hence, following these preventive measures properly along with vaccination is more effective to reduce the vulnerability of the population as well as the morbidity and mortality rates for building disaster resilient society (4-7).

The Nepal government in collaboration with other stakeholders, are conducting various activities such as preparation of Information, Education, Communication (IEC) material, and disseminate information to spread knowledge about COVID-19, through various medium such as social media, TV, radio, and mobile phone but the dissemination of knowledge to the community are still not satisfactory. Many of the marginalized and poor communities do not have access to these sources (8-11). In addition, the people having access to these sources of media are also not able to choose the authentic and correct information due to the lack of digital literacy as much of the misleading information on COVID-19 is also circulated through these sources (12). The absence of proper coordination, well-planned, and clearly defined roles and responsibilities of 3 tiers of government (local, provincial, and federal) regarding the COVID-19 pandemic have also affected the timely and efficient implementation of preventive measures (13). Lack of timely information to improve knowledge and preventive behavior on dealing with the crisis during a pandemic can further create anxiety and fear of being discredited and treated unfairly. Consequently, the general public might be reluctant to get tested which further delays the treatment with a large number of populations remaining undiagnosed (14, 15). Therefore, early identification of symptoms and timely treatment are very important in this scenario to control the spread of disease and fast recovery.

The lengthening of the pandemic might result in unprecedented challenges such as economic disruption, loss of jobs and business, and an increase in domestic violence among the general population (16-18). This would ultimately affect physical health such as discomfort, pain, lack of physical activities, and a sedentary lifestyle leads to weight gain and an increase the risk of cardiovascular disease, mental health such as depression as well as spiritual health (19-22). The large number of mutations and emergence of several variants of the concern (VOCs) of SARS CoV-2 is highly probable in the future as well with increased transmissibility, damaging effect on human health, and increased risk of hospitalization (23). In such circumstances, Nepal’s health care system is fragile to handle an extensive increase in morbidity rates during the pandemic. Also, delivering quality health care service is very tough and challenging compared to other developed countries due to the limited resources such as skilled human resources, infrastructures, and equipment. Although, the total health budget was increased by Nepal Government during pandemic in fiscal years 2020/21, 2021/22, the decrease in allocation of budget to hospitals and academics compared to previous fiscal years have aggravated the situation (24-26). In addition, the occurrence of high comorbidities further intensifies the complications resulting in high mortality (27-29).

In such situations, it is very important to understand what is known, what is believed and accepted, and what is done regarding the preventive measures concerning COVID-19. The necessity of a high level of psychological readiness i.e., belief and acceptance towards the severity of disease particularly for behavioral change (30) increased the importance of the study. In addition, previous studies conducted on Ethiopia (31, 32), Bangladesh (33, 34), Turkey (35), China (36, 37), Malaysia (38), Iran (39), Indonesia (40) and Cameroon (41) regarding knowledge, attitude and practice (KAP) on COVID-19 also provided substantiation of clear understanding of these concerns further underscored the significance of the study. However, there are limited studies regarding KAP on COVID-19 among general population in context of Nepal. Therefore, our study aims to observe the situation of knowledge, attitude and practice (KAP) on COVID-19 among the general population, and its relationship with socio-demographic characteristics. The study results will help to comprehend the knowledge gap and behavioral patterns that are important to prioritize the policy and allocate the resources for the implementation of intervention programs to address the challenges of the COVID-19 pandemic.

## 2. Methodology

### 2.1 Study area and sampling

The cross-sectional study was conducted among the general population above the age of 18 years from eight districts-Kathmandu, Bhaktapur, Lalitpur, Morang, Sunsari, Rupandehi, Chitwan, and Kaski of Nepal (Figure 1). A descriptive study design was used for analyzing the survey data. A convenient non-probability sampling method with a total sample size of 702 was considered due to the inappropriateness of the probability sampling during the period of the probability sampling during the period of the pandemic.

**Fig 1.** Districts selected for sampling. The number of samples in each of the eight selected districts was determined according to the proportion of the population of the districts. The sample size for each district was calculated as follows: Proportion of population in each district (A) = (Population of a district)/ (Total population of all eight districts) ×100%.

Sample size of each district (n) = (A)/100×Total sample size (702).

Hence, the number of participants selected in each district were Kathmandu-197, Bhaktapur-35, Lalitpur-5, Morang-107, Sunsari-88, Rupandehi-99, Chitwan-66, and Kaski-56.

### 2.2 Data collection

For data collection, we used a mixed method approach which included phone survey and face to face interview as it was difficult to interact and collect all the samples employing only one approach during the pandemics. Skilled volunteers associated with Nepal Red Cross Society (NRCS) district chapters were mobilized to conduct surveys.

### 2.3 Ethical concern

The study was approved by the Ethical Review Board of the Nepal Health Research Council. The verbal and written Informed consent were taken from all the respondents who participated in the study through telephone survey and face to face interview respectively.

### 2.4 Measurements

#### Knowledge, attitude, and practice (KAP)

Knowledge level was measured using binary questions (yes/ no). Knowledge of symptoms, preventive measures, quarantine, and overall knowledge was assessed using a different number of questions. Thirteen questions assessed the knowledge of ‘symptoms’ with a total score range of 0-13. The knowledge of symptoms was dichotomized with a cutoff value of 10 as those having good knowledge (>10) and poor knowledge (≤10). There were six questions related to knowledge on ‘preventive measures’ with total score in the range 0-6. The knowledge on preventive measures was dichotomized with cutoff value of 6 as having good and poor knowledge. Similarly, five questions related to knowledge on ‘precondition to stay in quarantine’ with the total score in the range of 0-5 were categorized as having good and poor knowledge with a cutoff value of 5. Lastly, there were twelve final questions to measure the ‘overall knowledge’ level of the general population ranging from 0-12 with a cutoff value of 7.

Attitude level was measured using a five-point Likert Scale questions where the responses varied from (1) strongly disagree to (5) strongly agree. The total score of the attitude questionnaire was the sum of scores of all the items ranging from 6 to 30 which was categorized into good and a bad attitude with a cutoff value of 26. Practice level was measured using a five-point Likert Scale questionnaire for prevention of COVID-19 from (1) never to (5) always. The total score of practice questionnaires ranging from 9 to 45 was categorized as good practice and poor practice with a cut-off value of 33.

#### Socio-demographic characteristics

The structured questionnaire was used to collect the data on socio-demographic characteristics such as geographical location (rural municipality, urban municipality), gender (male, female), age (<20 years, 20-30 years, 31-40 years, 41-50 years, >50 years), marital status (married, unmarried), education (no education, literate, basic education, secondary education, undergraduate, graduate and above), income (<NPR 5000, NPR 5000-10,000, NPR 10,000-15,000, NPR 15,000-20,000, NPR >20,000), occupation of the respondent (white-collar occupation - service, business, house rent; blue-collar occupation-agriculture, labor; and others - self-employed, remittance and others).

### 2.5 Statistical analysis

Descriptive statistics were used to calculate frequencies, proportions, and averages. Chi-square test was used to find the relationship between categorical variables. Similarly, the Wilcoxon Rank Test was used for continuous variables. The reliability of the questionnaire used to measure knowledge, attitude, and practice was evaluated using Cronbach’s Alpha. The alpha value of 0.6 is considered as acceptable. A multivariate logistic regression model was used to find the relationship of KAP with socio-demographic characteristics. Statistical analyses were conducted using Statistical Package for the Social Sciences V.19 (SPSS Inc, Chicago, Illinois, USA).

## 3. Results

### 3.1 Knowledge

The study revealed that, very few proportions of respondents had knowledge of the symptoms of diarrhea (n = 258, 37%), conjunctivitis (n = 159, 22.9%), skin rashes or discoloration of skin or toes (n = 172, 24.7%), loss of speech or movement (n = 313, 45%) whereas, a higher proportion of respondents had knowledge on fever (n = 699, 99.7%), dry cough 680, 97%), tiredness (n = 634, 90.3%), aches and pains (n = 630, 89.9%), sore throat (n = 622, 88.6%), headache (n = 620, 88.8%), loss of taste and smell (n = 651, 92.9%), difficulty breathing and shortness of breath (n = 654, 93.4%), and chest pain (n = 557, 80%) (Figure 2).

**Fig 2.** Knowledge of symptoms of the COVID-19 pandemic among the respondents. Regarding the preventive measures, a higher proportion of respondents showed good knowledge of the necessity of washing hands or use of alcohol-based sanitizer (n = 692, 98.6%), covering of nose and mouth while sneezing and coughing (n = 700, 99.7%), maintaining social distance (n = 683, 97.3%), use of mask (n = 698, 99.4%), avoiding crowd (n = 691, 98.4%), and staying home (n = 677, 96.6%) (Figure 3).

**Fig 3.** Knowledge of prevention on COVID-19 among the respondents. Similarly, higher proportion of respondents had knowledge of pre-conditions to stay in quarantine such as contact with droplets while sneezing and coughing 675 (96.4%) followed by taking care of an infected person within house (n = 649, 93%) and using utensils of infected person (n = 630, 90.4%) (Figure 4).

**Fig 4.** Knowledge of pre-condition to stay in quarantine among the respondents. Considering the knowledge of comorbidities during the COVID-19 pandemic, many of the respondents had knowledge of asthma (n = 568, 81.3%), followed by cardiovascular symptoms (n = 511, 73%), while very few had knowledge of cerebrovascular disease (n = 292, 41.8%) (Figure 5).

**Fig 5.** Knowledge of comorbidities during the COVID-19 pandemic among the respondents.

Table 1 comprising the overall knowledge on COVID-19 further revealed that a major proportion of respondents (n = 655, 93.3%) had good knowledge about overall preventive measures regarding COVID-19. However, very few respondents had overall knowledge of symptoms (n = 221, 32%) and comorbidities (n = 283, 40.7%). Similarly, (n = 466, 67.1%) of respondents had overall knowledge of the pre-condition of quarantine. Concerning the knowledge of survival of SARS CoV-2 in fomites, (n = 375, 53.6%) knew about its survival duration on plastic and stainless steel, (n = 280, 40%) on cardboard, (n = 306, 43.7%) on copper, and (n = 352, 50.2%) on aerosols.

**Table 1.** Knowledge level of respondents on COVID-19.

| S.N. | Items | Yes (%) |
| --- | --- | --- |
| K1 | Do you have good knowledge of the overall symptoms of COVID-19 as mentioned by WHO? | 221 (32%) |
| K2 | Do you have good knowledge of preventive measures to be protected from COVID-19? | 655 (93.3%) |
| K3 | Do you have good knowledge of pre-conditions to stay in quarantine? | 466 (67.1%) |
| K4 | Do you know a proper diet and exercise to boost the immune system? | 653 (93.4%) |
| K5 | Do you know SARS-CoV-2 can transfer from aerosols? | 429 (61.5%) |
| K6 | Do you know SARS-CoV-2 will stay alive for seventy-two hours in plastic, and stainless steel? | 375 (53.6%) |
| K7 | Do you know SARS-CoV-2 will stay alive for twenty-four hours in cardboard? | 280 (40%) |
| K8 | Do you know SARS-CoV-2 will stay alive for four hours in copper? | 306 (43.7%) |
| K9 | Do you know SARS-CoV-2 will stay alive for three hours in aerosols? | 352 (50.2%) |
| K10 | Do you know that the SARS-CoV-2 virus on the surface of mobile phones mostly comes into contact with the nose and ear? | 355 (51.2%) |
| K11 | Do you know about the new variants of SARS-CoV-2? | 421 (60.5%) |
| K12 | Do you have proper knowledge regarding several chronic diseases that increase the death rate of COVID-19 infected persons? | 283 (40.7%) |

### 3.2 Attitude

Regarding attitude, a significant proportion of respondents (n = 473, 67.4%) strongly agreed and (n = 199, 28.8%) agreed that keeping themselves safe was breaking the chain of transmission of COVID-19 and (n = 424, 61.2%) strongly agreed and (n = 214, 30.9%) agreed that exercise and diet would increase the immunity. Similarly, (n = 293, 41.7%) respondents strongly agreed and (n = 320, 45.6%) agreed that preventive measures would reduce the risk of transmission; (n = 338, 48.1%) strongly agreed and (n = 296, 42.2%) agreed that staying in quarantine would reduce the transmission and (n = 272, 39.1%) strongly agreed and (n = 353, 50.7%) agreed that pandemic could further lead to the difficult situation if not taken care. However, a very small proportion of respondents (n = 99, 14.3%) strongly agreed and (n = 215, 31.1%) agreed that they would soon get rid of the COVID-19 pandemic (Table 2).

**Table 2.** An attitude of respondents on COVID-19.

| S.N<br>. | Items | Strongly<br>agree | Agree | Neutral | Disagree | Strongly<br>disagree |
| --- | --- | --- | --- | --- | --- | --- |
| A1 | Do you think preventive measures will reduce the risk of transmission? | 293 (41.7%) | 320 (45.6%) | 82 (11.7%) | 3 (0.4%) | 4 (0.6%) |
| A2 | Do you think staying in quarantine will reduce the transmission? | 338 (48.1%) | 296 (42.2%) | 63 (9%) | 4 (0.6%) | 1 (0.1%) |
| A3 | Do you agree that keeping yourself safe is breaking the chain of transmission of COVID-19? | 473 (67.4%) | 199 (28.8%) | 24 (3.4%) | 4 (0.6%) | 2 (0.3%) |
| A4 | Do you believe that exercise and diet will increase immunity? | 424 (61.2%) | 214 (30.9%) | 41 (5.9%) | 5 (0.7%) | 9 (1.3%) |
| A5 | Do you believe that it can further lead to a difficult situation if not taken care of? | 272 (39.1%) | 353 (50.7%) | 59 (8.5%) | 9 (1.3%) | 3 (0.4%) |
| A6 | Do you believe we will soon get rid of the COVID-19 pandemic? | 99 (14.3%) | 215 (31.1%) | 282 (40.8%) | 56 (8.1%) | 39 (5.6%) |

### 3.3 Practice

During the lockdown, a higher proportion of respondents (n = 601, 86.1%) always used a mask while going outside followed by those who always covered their mouth and nose while coughing and sneezing (n = 576, 82.5%) and always washed their hands with soap for 20 sec or use alcohol-based sanitizer (n = 522, 74.6%). Nevertheless, very few respondents did regular exercise (n = 222, 31.7%) and cleaned mobile phones with sanitizer after coming from outside (n = 225, 32.2%) (Table 3).

**Table 3.** The practice of prevention among respondents during lockdown.

| S.N. | Items | Always | Most of the time | Sometime | Rarely | Never |
| --- | --- | --- | --- | --- | --- | --- |
| PL1 | Washing hands with soap for 20 sec or use alcohol-based sanitizer | 522 (74.6%) | 139 (19.9%) | 26 (3.7%) | 8 (1.1%) | 5 (0.7%) |
| PL2 | Cover mouth and nose while coughing and sneezing. | 576 (82.5%) | 98 (14%) | 15 (2.1%) | 5 (0.7%) | 4 (0.6%) |
| PL3 | Maintain social distance up to 6ft | 469 (67%) | 177 (25.3%) | 37 (5.3%) | 12 (1.7%) | 5 (0.7%) |
| PL4 | Use a mask while going outside | 601 (86.1%) | 72 (10.3%) | 15 (2.1%) | 6 (0.9%) | 4 (0.6%) |
| PL5 | Avoid crowd | 466 (66.7%) | 183 (26.2%) | 25 (3.6%) | 19 (2.7%) | 6 (0.9%) |
| PL6 | Stay at home for safety | 502 (71.7%) | 149 (21.3%) | 20 (2.9%) | 22 (3.1%) | 7 (1%) |
| PL7 | Clean mobile phone with sanitizer after coming from outside | 225 (32.2%) | 108 (15.5%) | 103 (14.7%) | 69 (9.9%) | 194 (27.8%) |
| PL8 | Having proper diet | 433 (61.9%) | 199 (28.5%) | 32 (4.6%) | 24 (3.4%) | 11 (1.6%) |
| PL9 | Doing regular exercise | 222 (31.7%) | 160 (22.9%) | 164 (23.4%) | 31 (4.4%) | 123 (17.6%) |

After lockdown, (n = 439, 63.6%) always covered the mouth and nose while coughing and sneezing. However, very few of them always stayed at home for safety (n = 53, 7.6%), cleaned mobile phones with sanitizer after coming from outside (n = 57, 8.2%), and avoided crowds (n = 58, 8.4%) (Table 4). The median score for practice for prevention during lockdown was 40 and after the lockdown was 33. The Wilcoxon Rank Test showed that the relationship was statistically significant (p<0.001).

**Table 4.**
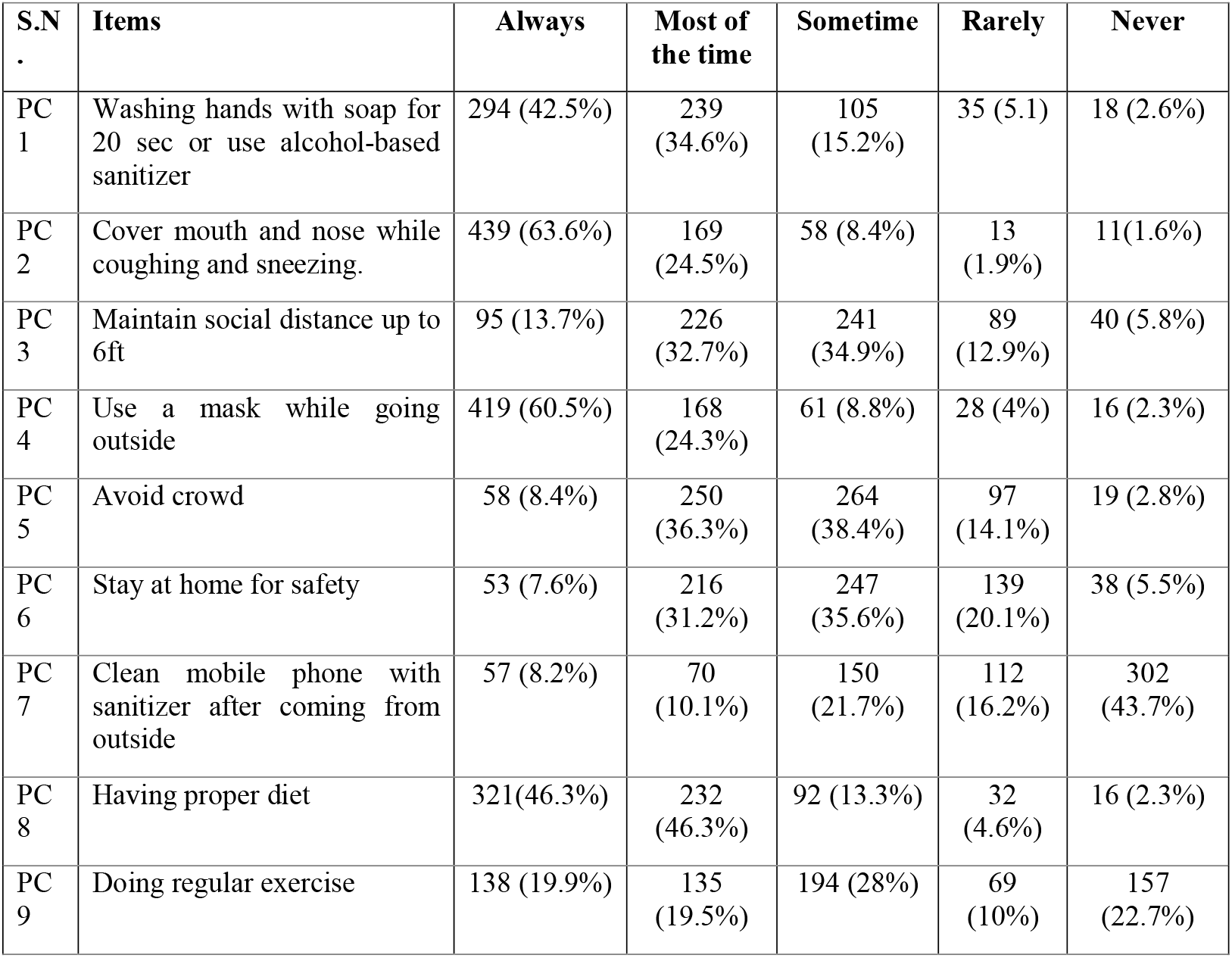
The practice for prevention among respondents after lockdown.

| S.N<br>. | Items | Always | Most of<br>the time | Sometime | Rarely | Never |
| --- | --- | --- | --- | --- | --- | --- |
| PC<br>1 | Washing hands with soap for 20 sec or use alcohol-based sanitizer | 294 (42.5%) | 239 (34.6%) | 105 (15.2%) | 35 (5.1) | 18 (2.6%) |
| PC<br>2 | Cover mouth and nose while coughing and sneezing. | 439 (63.6%) | 169 (24.5%) | 58 (8.4%) | 13 (1.9%) | 11(1.6%) |
| PC<br>3 | Maintain social distance up to 6ft | 95 (13.7%) | 226 (32.7%) | 241 (34.9%) | 89 (12.9%) | 40 (5.8%) |
| PC<br>4 | Use a mask while going outside | 419 (60.5%) | 168 (24.3%) | 61 (8.8%) | 28 (4%) | 16 (2.3%) |
| PC<br>5 | Avoid crowd | 58 (8.4%) | 250 (36.3%) | 264 (38.4%) | 97 (14.1%) | 19 (2.8%) |
| PC<br>6 | Stay at home for safety | 53 (7.6%) | 216 (31.2%) | 247 (35.6%) | 139 (20.1%) | 38 (5.5%) |
| PC<br>7 | Clean mobile phone with sanitizer after coming from outside | 57 (8.2%) | 70 (10.1%) | 150 (21.7%) | 112 (16.2%) | 302 (43.7%) |
| PC<br>8 | Having proper diet | 321(46.3%) | 232 (46.3%) | 92 (13.3%) | 32 (4.6%) | 16 (2.3%) |
| PC<br>9 | Doing regular exercise | 138 (19.9%) | 135 (19.5%) | 194 (28%) | 69 (10%) | 157 (22.7%) |

### 3.4 Relationship of KAP and socio-demographic characteristics

Cronbach’s alpha (α) value of the questionnaire for measuring knowledge level and attitude on COVID-19 was 0.8 and 0.6. Similarly, α value of questionnaire practice for prevention on COVID-19 during and after the lockdown was 0.8. The good knowledge on COVID −19 was significantly higher among the residents of urban areas (n = 288, 48.5%) (p<0.001), unmarried (n = 113, 52.8%) (p<0.05), following white-collar occupation 205 (51.8%) (p<0.001), educational level with graduate and above 49 (75.4%) (p<0.001) and high-income level 252 (58.3%) (p<0.001).

A good attitude was significantly higher among the unmarried respondents (n = 86, 40.8%) (p<0.01), those following the white-collar occupation (n = 160, 39.4%) (p<0.001), and high-income group (n = 166, 37.4%) (p<0.001). In the same way, good practice for prevention on COVID −19 was significantly higher among the respondents from urban areas (n = 288, 46.7%) (p<0.01), unmarried group (n = 109, 51.2%) (p<0.05), following white-collar occupation (n = 220, 54.1%) (p<0.001), high-educated group (n = 42, 60.9%) (p<0.001) and having high-income (n = 245, 55.2%) (p<0.001 (Table 5).

**Table 5.** Socio-demographic characteristics of the population with different levels of KAP on COVID-19.

| Variables | Knowledge |  | Chi-square test (p-value) | Attitude |  | Chi-square test (p-value) | Practice |  | Chi-square test (p-value) |
| --- | --- | --- | --- | --- | --- | --- | --- | --- | --- |
|  | Number of respondents | Good Knowledge (%) |  | Number of respondents | Good Attitude (%) |  | Number of respondents | Good Practice (%) |  |
| **Municipality** |  |  |  |  |  |  |  |  |  |
| Rural | 62 | 15 (24.2%) |  | 65 | 14 (22.6%) | 0.07 | 60 | 16 (26.7%) | .003 |
| Urban | 594 | 288 (48.5%) | <0.001 | 631 | 207 (33.7%) |  | 617 | 288 (46.7%) |  |
| **Gender** |  |  |  |  |  |  |  |  |  |
| Male | 312 | 150 (48.9%) | 0.45 | 318 | 93 (29.2%) | 0.09 | 326 | 138 (42.3%) | 0.14 |
| Female | 350 | 158 (45.1%) |  | 364 | 128 (35.2%) |  | 357 | 171 (47.9%) |  |
| **Marital status** |  |  |  |  |  |  |  |  |  |
| Married | 446 | 193 (43.3%) | 0.02 | 469 | 135 (28.8%) | 0.002 | 468 | 199 (42.5%) | 0.03 |
| Unmarried | 214 | 113 (52.8%) |  | 211 | 86 (40.8%) |  | 213 | 109 (51.2%) |  |
| **Main occupation** |  |  |  |  |  |  |  |  |  |
| Blue-collar | 115 | 30 (26.1%) | <0.001 | 120 | 17 (14.2%) | <0.001 | 118 | 28 (23.7%) | <0.001 |
| White-collar | 396 | 205 (51.8%) |  | 406 | 160 (39.4%) |  | 407 | 220 (54.1%) |  |
| Others | 118 | 58 (49.2%) |  | 123 | 36 (29.3%) |  | 123 | 46 (37.4%) |  |
| **Age (years)** |  |  |  |  |  |  |  |  |  |
| <20 | 32 | 9 (28.1%) | 0.02 | 29 | 9 (31%) | 0.007 | 33 | 15 (45.5%) | 0.41 |
| 20-30 | 256 | 134 (52.3%) |  | 256 | 98 (38.3%) |  | 251 | 117 (46.6%) |  |
| 31-40 | 153 | 71 (46.4%) |  | 156 | 57 (36.5%) |  | 157 | 63 (40.1%) |  |
| 41-50 | 120 | 55 (45.8%) |  | 132 | 33 (25%) |  | 134 | 68 (50.7%) |  |
| >50 | 99 | 37 (37.4%) |  | 106 | 23 (21.7%) |  | 105 | 44 (41.9%) |  |
| **Education of respondent** |  |  |  |  |  |  |  |  |  |
| No Education | 49 | 7 (14.3%) |  | 51 | 11 (21.6%) | 0.09 | 54 | 8 (14.8%) | <0.001 |
| Literate | 19 | 3(15.8%) | <0.001 | 23 | 6 (26.1%) |  | 22 | 8 (36.4%) |  |
| Basic Education | 83 | 19 (22.9%) |  | 87 | 20 (23%) |  | 87 | 24 (27.6%) |  |
| Secondary Education | 220 | 93 (42.3%) |  | 225 | 74 (32.8%) |  | 221 | 94 (42.5%) |  |
| Undergraduate | 224 | 135 (60.3%) |  | 225 | 84 (37.3%) |  | 227 | 130 (57.3%) |  |
| Graduate and above | 65 | 49 (75.4%) |  | 68 | 24 (35.3%) |  | 69 | 42 (60.9%) |  |
| **Income- Nepalese Rupee (NPR)** |  |  |  |  |  |  |  |  |  |
| <5000 | 25 | 6 (24%) | <0.001 | 28 | 3 (10.7%) | <0.001 | 27 | 5 (18.5%) | <0.001 |
| 5000-10,000 | 48 | 8 (16.7%) |  | 47 | 7 (14.9%) |  | 48 | 10 (20.8%) |  |
| 10,000-15,000 | 54 | 9 (16.7%) |  | 60 | 11 (18.3%) |  | 59 | 11 (18.6%) |  |
| 15,000-20,000 | 100 | 32 (32%) |  | 100 | 34 (34%) |  | 102 | 38 (37.3%) |  |
| >20,000 | 432 | 252 (58.3%) |  | 444 | 166 (37.4%) |  | 444 | 245 (55.2%) |  |

Logistic regression analysis showed that the respondents in urban areas were 2.34 times more likely to have knowledge (95% CI 1.19 to 5.39) than those staying in rural areas. Similarly, those with white-collar (OR: 1.24; 95% CI 0.70 to 2.19) and other occupations (OR 1.68, 95% CI 0.88 to 3.19) had higher odds of having knowledge compared to blue-collar occupations. Highly educated respondents such as graduate and above (OR: 11.54; CI 3.58 to 38.10), undergraduate (OR: 6.90; CI 2.44 to 37.23), having secondary education (OR: 4.54; 95% CI 1.66 to 12.34) had higher odds of having knowledge than the non-educated group. Likewise, the odds ratio of having good knowledge on prevention among the higher income group with monthly income >NRs 20,000 was 2.57 (95% CI 0.84 to 7.90).

Regarding attitude, people with white-collar occupations (OR: 3.08, 95% CI 1.67 to 5.68) and other occupations (OR: 1.98, 95% CI 0.77 to 5.04) had high odds of having a good attitude compared to those having blue-collar occupation.

In the same way, urban people were 1.99 times more likely to adopt the good practice for prevention (95% CI 1.04 to 3.79) compared to rural people. Also, females were 1.70 times more likely to have good preventive practice (95% CI 1.19 to 2.43) than males. Our study further revealed that the odds ratio of practicing good preventive behavior was 1.79 times (95% CI 1.04 to 3.08) for those with white-collar occupations. Moreover, educated respondents such as graduate and above (OR: 7.82, 95% CI 2.66 to 22.93), undergraduate (OR: 7.53, 95% CI 2.84 to 20.01), secondary education (OR: 5.52, 95% CI 2.15 to 14.12) and basic education (OR: 3.06, 95% CI 1.13 to 8.29) had higher odds of practicing good preventive behavior compared to non-educated group (Table 6).

**Table 6.** Odds ratios of having good knowledge, attitude, practice on COVID-19 among the general population according to the socio-demographic characteristics.

| Variables | Good knowledge |  | Good attitude |  | Good practice |  |
| --- | --- | --- | --- | --- | --- | --- |
|  | Univariate | Multivariate | Univariate | Multivariate | Univariate | Multivariate |
| **Municipality** |  |  |  |  |  |  |
| Rural | 1 | 1 | 1 | 1 | 1 | 1 |
| Urban | 2.94 (1.61 to 5.39)*** | 2.34 (1.19 to 4.57)* | 1.73 (0.93 to 3.22) | 1.72 (0.87 to 3.40) | 2.40 (1.33 to 4.35)** | 1.99 (1.04 to 3.79)* |
| **Gender** |  |  |  |  |  |  |
| Male | 1 | 1 | 1 | 1 | 1 | 1 |
| Female | 0.88 (0.65 to 1.20) | 0.96 (0.67 to 1.41) | 1.32 (0.94 to 1.81) | 1.32 (0.90 to 1.90) | 1.25 (0.92 to 1.69)** | 1.70 (1.19 to 2.43)** |
| **Marital status** |  |  |  |  |  |  |
| Unmarried | 1 | 1 | 1 | 1 | 1 | 1 |
| Married | 0.68 (0.49 to 0.94)* | 0.89 (0.52 to 1.52) | 0.58 (0.41 to 0.82)** | 0.63 (0.37 to 1.04) | 0.70 (0.51 to 0.97)* | 0.61 (0.35 to 1.08) |
| **Main occupation** |  |  |  |  |  |  |
| Blue-collar | 1 | 1 | 1 | 1 | 1 | 1 |
| White-collar | 3.04 (1.91 to 4.82)*** | 1.24 (0.70 to 2.19)*** | 3.94 (2.27 to 6.83)*** | 3.08 (1.67 to 5.68) *** | 3.78 (2.37 to 6.03)*** | 1.79 (1.04 to 3.08)* |
| Others | 2.73 (1.57 to 4.75)*** | 1.68 (0.88 to 3.19)** | 2.50 (1.31 to 4.77)** | 1.98 (0.77 to 5.04)* | 1.92 (1.09 to 3.36)* | 1.13 (0.60 to 2.12) |
| **Age (years)** |  |  |  |  |  |  |
| <20 | 1 | 1 |  | 1 | 1 | 1 |
| 20-30 | 2.80 (1.25 to 6.30)* | 3.46 (1.32 to 9.03)** | 1.37 (0.60 to 3.14) | 1.98 (0.77 to 5.04) | 1.04 (0.50 to 2.17) | 1.18 (0.50 to 2.78) |
| 31-40 | 2.21 (0.96 to 5.03) | 3.89 (1.30 to 11.63)* | 1.27 (0.54 to 2.99) | 2.36 (0.81 to 6.84) | 0.80 (0.37 to 1.71) | 1.48 (0.54 to 4.01) |
| 41-50 | 2.16 (0.92 to 5.06) | 4.06 (1.29 to 12.74)* | 0.74 (0.30 to 1.78) | 1.36 (0.44 to 4.17) | 1.23 (0.57 to 2.65) | 2.66 (0.93 to 7.56) |
| >50 | 1.52 (0.63 to 3.64) | 4.81 (1.47 to 15.68)** | 0.61 (0.24 to 1.53) | 1.13 (0.35 to 3.64) | 0.86 (0.39 to 1.90) | 2.71 (0.91 to 8.03) |
| **Education of respondent** |  |  |  |  |  |  |
| No Education | 1 | 1 | 1 | 1 | 1 | 1 |
| Literate | 1.12 (0.25 to 4.89) | 0.90 (0.18 to 4.40) | 1.28 (0.40 to 4.03) | 0.86 (0.23 to 3.20) | 3.28 (1.04 to 10.35)* | 3.33 (0.92 to 12.01) |
| Basic Education | 1.78 (0.68 to 4.60) | 1.87 (0.63 to 5.49) | 1.08 (0.47 to 2.49) | 0.84 (0.31 to 2.16) | 2.19 (0.90 to 5.31) | 3.06 (1.13 to 8.29)* |
| Secondary Education | 4.39 (1.89 to 10.21)** | 4.54 (1.66 to 12.34)** | 1.78 (0.86 to 3.67) | 1.04 (0.44 to 2.46) | 4.25 (1.91 to 9.44)*** | 5.52 (2.15 to 14.12)*** |
| Undergraduate | 9.10 (3.91 to 21.16)*** | 6.90 (2.44 to 37.23)*** | 2.16 (1.05 to 4.45) | 0.76 (0.30 to 1.89) | 7.70 (3.47 to 17.04)*** | 7.53 (2.84 to 20.01)*** |
| Graduate and above | 18.37 (6.90 to 48.91)*** | 11.54 (3.58 to 38.10)*** | 1.98 (0.86 to 4.55) | 0.56 (0.20 to 1.58) | 8.94 (3.66 to 21.84)*** | 7.82 (2.66 to 22.93)*** |
| **Income-Nepalese Rupees (NPR)** |  |  |  |  |  |  |
| <5000 | 1 | 1 | 1 | 1 | 1 | 1 |
| 5000-10,000 | 0.63 (0.19 to 2.08) | 0.93 (0.24 to 3.55) | 1.45 (0.34 to 6.16) | 2.03 (0.37 to 11.06) | 1.15 (0.35 to 3.82) | 1.29 (0.33 to 5.02) |
| 10,000-15,000 | 0.63 (0.19 to 2.02) | 0.55 (0.14 to 2.14) | 1.87 (0.47 to 7.32) | 2.04 (0.40 to 10.39) | 1 .008 (0.31 to 3.25) | 0.94 (0.25 to 3.50) |
| 15,000-20,000 | 1.49 (0.54 to 4.08) | 1.41 (0.43 to 4.57) | 4.29 (1.20 to 15.24)* | 4.76 (1.01 to 22.38) * | 2.61 (0.91 to 7.47) | 2.19 (0.65 to 7.35) |
| >20,000 | 4.43 (1.73 to 11.32)** | 2.57 (0.84 to 7.90)* | 4.97 (1.48 to 16.73)* | 4.29 (0.94 to 19.54) | 5.47 (2.01 to 14.56)*** | 2.62 (0.82 to 8.35) |

## 4. Discussion

Our study aimed to understand the knowledge, attitude, and practice (KAP on COVID-19 pandemic and its relationship with socio-demographic characteristics among the general population in Nepal.

The higher proportion of respondents had good knowledge of symptoms such as fever, dry cough, tiredness, aches and pains, sore throat, loss of taste or smell, difficulty breathing or shortness of breath and very few had knowledge of symptoms such as diarrhea, conjunctivitis, rash on the skin, or discoloration of fingers or toes and loss of speech or movement. The knowledge of the overall symptoms of COVID-19 among the community was observed to be very low although the understanding is very essential during the pandemic. Other similar studies also revealed that a higher proportion of community people had knowledge of symptoms such as fever and cough and very less knew about the other symptoms of COVID-19 compared to our study (31, 33). A lower level of knowledge on overall symptoms might prevent the early detection of disease and hinder the timely treatment (15).

Our study illustrated that knowledge on preventive behaviors seemed to be high which is in line with similar study conducted in Bangladesh (33). However, the knowledge of survival period of SARS-CoV-2 in an external environment was low, which might increase the risk of disease transmission. Similarly, the low knowledge associated with comorbidities such as diabetes, Chronic obstructive Pulmonary Disease, hypertension, malignancy, etc. among the study population can further augment the severity of COVID-19 patients leading to hospitalization, admission to intensive care unit, and increase in the mortality rate (42).

Our study further showed that the majority of people had a positive attitude such that most of them agree that preventive measures will reduce the risk of transmission, pre-conditions required to stay in quarantine, keeping themselves safe, proper diet and exercise, and taking necessary steps to reduce possible difficulties. However, they were not convinced that they would get rid of the pandemic soon. After the lockdown, the preventive practice is observed to be lowered compared to the practice during the lockdown. The reduction in the mortality rate does not mean that the pandemic will end soon. There is always the risk of the emergence of new VOCs that might be responsible for new waves of the COVID-19 pandemic. The negligence of community people was observed though they had knowledge and information. The result was in line with the study conducted in Janakpur, Nepal which revealed that though the perception on COVID-19 was not so different, the preventive behaviors were reduced in post-lockdown compared to the behaviors during the lockdown. The less precautionary behavior was especially observed among those following unofficial sources of information after lockdown compared to those following official government sites (43). During a humanitarian crisis, such as a pandemic, the spread of rumors, and misleading information through social media and various other communication mediums especially unofficial sources can further impact the KAP level of community people which led to an increase in morbidity and mortality (44). Such misinformation might affect the psychology and behavioral patterns of people hindering even simple practices such as hand washing, and social distancing (45). Therefore, it is important to trace the source of misinformation and make people aware of its impact (46).

The regular monitoring and study regarding KAP on COVID-19 during the pandemic should be prioritized by concerned authorities.

Our study uncovered that the KAP on COVID-19 among the general population were influenced by socio-demographic characteristics. The population living in the urban area had good knowledge on COVID-19 and good practice in prevention compared to those living in rural areas. The findings were similar to the study conducted in Bangladesh (33, 34). The reason behind this could be that the people living in rural areas do not have easy access to information sources. In contrast, urban people have easy access to various types of information sources that increase their familiarity on COVID-19 (47). Besides, the protection facilities are easily available in these areas to adopt preventive measures.

The respondents with white-collar occupations had good KAP. The result was in line with the study conducted in Ethiopia which showed that the people working as a government officers and merchant were more likely to have sufficient knowledge on COVID-19 than those working as a laborer (47). Similar studies conducted in Turkey (35) and China (36) also illustrated the higher knowledge level on COVID-19 among the white-collar workers. The respondents having white-collar occupations mostly have easy access to information and strong learning ability further aids in increasing their attitude and practice. Our study further revealed that the highly educated respondents had good KAP on COVID-19; and this bear a resemblance to the study conducted in North East Ethiopia (32) and Bangladesh (33, 34). Highly educated people are more sensitive and have a better understanding of pertinent issues in society. Accordingly, they perceive the impact of pandemics more seriously, improving their attitude and practice for prevention along with the knowledge received (37).

Similarly, high-income respondents were observed to have good KAP in our study recapitulating the studies conducted in Malaysia (38) and Bangladesh (34). High-income people are mostly educated and low-income people are least educated with lower understanding. Furthermore, females were more likely to practice preventive behavior compared to males. The finding was similar to the studies conducted in Iran (39) and Bangladesh (33, 34). This result can be supported by the fact that women are more concerned about their health because they are socialized to be more serious and responsible about themselves and their family members’ health issues (40, 48, 49).

Knowledge itself is the foundation of learning, and the basis of establishing attitude, and attitude further influence the practice of people (36). Our study showed that the KAP were different according to the varying socio-demographic conditions. Moreover, the differences in learning abilities according to the diverse socio-demographic characteristics among the Nepalese population were highlighted in previous literatures (50, 51).. The pace of obtaining knowledge, and developing the attitude and practice of an individual depends upon their intellectual ability and previous knowledge (52). To reduce this gap among people of different socio-demographic statuses and to make communication more effective, it is important to identify the differences between messages disseminated and received as a result of differential exposure to the intervention, inconsistencies in interpretation, and transforming information (53). It is important to consider the channel through which the message is disseminated, to whom the message is attributed, the response of the audience, and the feature of the message.

Ministry of Health and Population, Nepal Government has prepared, ‘Guideline for Mobilization of COVID-19 Facilitation Group in Community Level’ (54), ‘Public Health Standards Regarding COVID-19 during Festivals and Celebrations’ (55), ‘COVID-19 Micro Contaminant Plan’ (56), and ‘Environmental Cleaning and Disinfection Interim Guidance in the context of COVID-19’ (57), which highlighted the prevention measures in households to community level and also the regular monitoring of preventive behavior among the community people.

The key messages highlighted in these documents should be translated to different languages and also in understandable form by the entire population including poor and marginalized people. Besides, the information disseminated should be based on scientific evidence to convince the people. Although the ‘Guideline on the Distribution of Isolation Kits for Home Isolation Cases’ (58) was prepared by Nepal Government, the distribution of kit consisting materials for preventive measures should also be emphasized especially among the most needful people such as those with a low-economic conditions or rural people who have poor behavioral practices in prevention.

A targeted program is necessary to improve the KAP on COVID-19 according to gender, occupation, income, educational background, and area of residence that will aid to control the risk of further waves of the pandemic in the future. Moreover, the use of social norms theory should be emphasized to accomplish positive health outcomes. Therefore, it is important to understand the approach to integrating social norms for better communication and improving the knowledge, attitude, and practice on COVID-19 (59, 60). The needful and effective steps by concerned stakeholders from diverse disciplines will help to aware the community and augment preventive behavior.

## 5. Conclusions

Findings from our study are imperative to understand the perceived knowledge, attitude and behavior during the COVID-19 pandemic according to the various socio-demographic characteristics. The knowledge level among the general population was good regarding symptoms, preventive practices, and pre-conditions to stay in quarantine. However, only a few of them knew about the survival duration of SARS-CoV-2 in fomites such as stainless steel, plastic, cardboard, copper, and aerosols. Regarding attitude, few people believed that they would soon get rid of the pandemic. Concerning practice for prevention, few people practiced staying at home safely, avoiding crowds, and cleaning mobile phones after coming from outside, while many of them frequently washed their hands with soap or used sanitizer, covered their mouth or nose while sneezing or coughing, used masks and took proper diet. The overall practice of prevention was reduced after the lockdown compared to the practice during the lockdown.

The people residing in urban areas, with white-collar occupations, high-income, and high-education were good at KAP on COVID-19. In addition, females seem to be good at practicing preventive behavior although their knowledge level was lower compared to male.

The KAP on COVID-19 were found to be different among the people with varying socio-economic conditions due to the difference in the level of their understanding and available resources. The study result recommends the subsequent improvement in existing plans, policies, and guidelines for their effective implementation considering effectual dissemination of information as well as infodemic management. The multifaceted approach along with the significant contribution of concerned stakeholders is essential to deal with the challenges during such a health crisis with effective utilization of the available limited resources.

## Data Availability

All relevant data are within the manuscript and its Supporting Information files.

## Acknowledgments

The authors would like to thank all the volunteers associated with Nepal Red Cross Society (NRCS) district chapters for their contribution in questionnaire survey and valuable suggestions during the survey; and all the respondents of the survey.

## Authors’ contribution

Conceptualization: Salina Shrestha, Rabin Malla, Sadhana Shrestha, Pallavi Singh, Jeevan B. Sherchand Data Curation: Salina Shrestha

Formal analysis: Salina Shrestha

Funding Acquisition: Rabin Malla

Investigation: Rabin Malla, Salina Shrestha, Pallavi Singh

Methodology: Salina Shrestha, Rabin Malla, Sadhana Shrestha, Pallavi Singh

Project Administration: Rabin Malla, Pallavi Singh, Salina Shrestha

Supervision: Jeevan B. Sherchand, Rabin Malla, Sadhana Shrestha, Pallavi Singh

Writing Original draft and preparation: Salina Shrestha, Rabin Malla

Writing-Review & Editing: Sadhana Shrestha, Jeevan B. Sherchand, Rabin Malla

## Data Availability Statement

Data will be available on request.

